# Validation of Genomic and Transcriptomic Models of Homologous Recombination Deficiency in a Real-World Pan-Cancer Cohort

**DOI:** 10.1101/2021.12.20.21267985

**Authors:** Benjamin Leibowitz, Bonnie V Dougherty, Joshua SK Bell, Joshuah Kapilivsky, Jackson Michuda, Andrew Sedgwick, Wesley Munson, Tushar Chandra, Jonathan R Dry, Nike Beaubier, Catherine Igartua, Timothy Taxter

## Abstract

**Background:** With the introduction of DNA-damaging therapies into standard of care cancer treatment, there is a growing need for predictive diagnostics assessing homologous recombination deficiency (HRD) status across tumor types. Following the strong clinical evidence for the utility of DNA-sequencing-based HRD testing in ovarian cancer, and growing evidence in breast cancer, we present analytical validation of the Tempus|HRD-DNA test. We further developed, validated, and explored the Tempus|HRD-RNA model, which uses gene expression data from 16,470 RNA-seq samples to predict HRD status from formalin-fixed paraffin-embedded (FFPE) tumor samples across numerous cancer types.

**Methods:** Genomic and transcriptomic profiling was performed using next-generation sequencing from Tempus|xT, Tempus|xO, Tempus|xE, Tempus|RS, and Tempus|RS.v2 assays on 48,843 samples. Samples were labeled based on their *BRCA1, BRCA2* and selected Homologous Recombination Repair (HRR) pathway gene (*CDK12, PALB2, RAD51B, RAD51C, RAD51D*) mutational status to train and validate HRD-DNA, a genome-wide loss-of-heterozygosity biomarker, and HRD-RNA, a logistic regression model trained on gene expression, using several performance metrics and statistical tests.

**Results:** In a sample of 2,058 breast and 1,216 ovarian tumors, BRCA status was predicted by HRD-DNA with F1-scores of 0.98 and 0.96, respectively. Across an independent set of 1,363 samples across solid tumor types, the HRD-RNA model was predictive of BRCA status in prostate, pancreatic, and non-small cell lung cancer, with F1-scores of 0.88, 0.69, and 0.62, respectively.

**Conclusions:** We predict HRD-positive patients across many cancer types and believe both HRD models may generalize to other mechanisms of HRD outside of BRCA loss. HRD-RNA complements DNA-based HRD detection methods, especially for indications with low prevalence of BRCA alterations.

## Introduction

Genomic instability is an enabling characteristic of cancer that is often mediated by deficiency in DNA damage sensing and repair processes (1). The homologous recombination repair (HRR) pathway, which is responsible for repairing DNA double-strand breaks (DSBs) (2), is frequently dysregulated in cancer, leading to the accumulation of genomic defects and cancer progression (3). *BRCA1* and *BRCA2* are foundational to the HRR pathway and were initially discovered due to their association with hereditary breast and ovarian cancers. Since their discovery, screening for germline BRCA alterations has become a powerful tool for clinical risk assessment and management (4). In addition, understanding the role of *BRCA1* and *BRCA2* in HRR has led to the development of targeted therapies, such as poly-ADP ribose polymerase (PARP) inhibitors. This class of drugs exploits synthetic lethality with the base excision repair pathway to directly target the underlying mechanism contributing to tumorigenesis (5,6).

Genetically or epigenetically driven loss of function in *BRCA1* or *BRCA2* is the canonical driver of the homologous recombination deficiency (HRD) phenotype, which is defined as the inability to repair DSBs with HRR (7). Nevertheless, multiple genes may impact the ability of the HRR pathway to repair DSBs. Specifically, *BRCA1/2* alterations are not necessary to cause the HRD phenotype; alterations in other HRR-related genes (i.e., *RAD51C* and *PALB2*) have also been associated with the HRD phenotype (8). However, the set of necessary and sufficient genetic and epigenetic alterations that drive the clinical manifestation of the HRD phenotype—and thus potential sensitivity to DNA damaging therapies (PARP inhibitors) in specific cancer indications—has yet to be comprehensively determined.

Additional biomarkers, outside of mutations in *BRCA1/2*, are needed to better characterize the HRD phenotype and to identify patients without *BRCA1/2* biallelic loss who are most likely to benefit from DNA repair-targeting therapies. One such biomarker is the presence of genomic scars which are created when HR-deficient cells are unable to repair DNA damage. Genome-wide loss-of-heterozygosity measures genomic scarring by calculating the percent of the profiled genome with loss of at least one allele. Genome-wide loss-of-heterozygosity (gwLOH) has demonstrated clinical benefit detecting HRD in ovarian cancers when used either independently (9) or in combination with measures of telomeric allelic imbalance and large-scale state transitions (10). Many of these DNA-based HRD biomarkers are measured over regions in the absence of aneuploidy, defined by loss or gains of chromosomes or chromosome arms, and is considered a confounding variable that may inflate gwLOH (9,11–13).

An orthogonal approach is to measure genomic scars via mutational signatures (8). Using whole-genome sequencing data, mutational signatures have suggested 20-30% more breast cancer patients may harbor HRD than what is detectable using the *BRCA1/2* genotype alone (14–16). These methods orthogonal to *BRCA1/2* status have only demonstrated utility in breast and ovarian cancer, necessitating new approaches for other cancer types. Alternative approaches to detect HRD include measurement of epigenetic silencing of *BRCA1/2 (17–19)* and/or epigenetic or genetic loss of other gene members of the HRR pathway (5,20,21). However, these mechanisms may be tissue specific, requiring additional research between specific alterations and the HRD phenotype. Further, reliance on fresh tissue for whole-genome sequencing and the measurement of multiple molecular modalities can be impractical and costly in real-world clinical practice.

Functional biomarkers of HRD, such as those assessing RAD51 nuclear localization (22), have received increasing attention given the accumulating evidence for mechanisms of resistance to platinum chemotherapy (23) and PARP inhibitors (24), suggesting that HRD is a dynamic phenotype. DNA-based biomarkers of the HRD phenotype have a strong temporal dependency given that a sufficient accumulation of genomic scars are needed for detection. Additionally, genomic markers of instability may have limited reversibility and could represent the molecular history of the tumor rather than the current state of HRR proficiency. In contrast to DNA-based approaches, gene expression has the potential to capture the dynamic state of HRD in a manner that is independent from genomic scarring. Transcriptional signatures have shown promise in predicting BRCA status or genomic scars in prostate and pancreatic cancer (25–27). However, it has yet to be demonstrated that a transcriptome-based model can generalize across solid tumors.

Here, we present an analytical validation of the Tempus|HRD platform comprising two assays: HRD-DNA, which measures gwLOH to predict HRD status in breast and ovarian cancers, and HRD-RNA, a logistic-regression model trained on whole-exome capture RNA sequencing data that predicts HRD status across all other solid tumors. Using data from a large-scale, real-world cohort, we demonstrate the capabilities of these models to accurately detect HRD driven by *BRCA1/2* loss and non-*BRCA1/2* mechanisms.

## Methods

### IRB

All analyses were performed using de-identified data; IRB exemption Pro00042950 was obtained from Advarra on April 15, 2020.

### Sample selection

Prior to sequencing, a hematoxylin and eosin (H&E) stained slide was prepared for FFPE tumor specimens and reviewed by a board-certified pathologist to ensure that adequate tissue, tumor content, and sufficient nucleated cells were present to satisfy the minimum tumor content requirement. A minimum tumor content of 20% was required to result in adequate yield at extraction and to proceed with sequencing. Macrodissection was carried out when deemed feasible by a pathologist to increase the tumor content of a specimen. Macrodissection was required if the tumor percentage was less than 40%, and was performed to increase tumor content in some instances.

Sample metadata, specifically tumor purity and cancer cohort labels, was determined by board-certified pathologists. Sample status (i.e. primary, metastatic, lymph node) was determined using a rule set based on heuristics between ICD-10 diagnosis codes and ICD-09 codes for anatomical biopsy site locations. Tissue sites provided by external pathology reports were mapped to ICD-09 codes. Samples that were unambiguously primary samples (e.g., ovarian cancer biopsied from the ovary) were labeled as “Primary”. Samples that were unambiguously metastatic samples (e.g., ovarian cancer biopsied from the liver) were labeled as “Metastatic”. Samples biopsied from a regional lymph node were labeled “Intermediate - Lymph Involvement”. Samples with incomplete biopsy site location information provided on an external pathology report were labeled “Intermediate - Missing Data”.

For orthogonal concordance testing between HRD-DNA and 1p FISH results, samples were ensured to be gliomas biopsied from the brain, with at least 40% tumor purity, and curated with positive or negative FISH results performed on chromosome 1p within 6 months of collection of the biopsy used for xT sequencing.

### DNA and RNA sequencing

A representative sample of de-identified records from 48,843 FFPE tumor samples across 42 solid tumor cancer types with DNA and RNA sequencing data were selected from the Tempus Oncology Database. All underwent DNA sequencing based on the Tempus|xT targeted panel (n = 48,827), Tempus|xO (n = 9), or Tempus|xE whole-exome panel (n = 17) these samples, 47,997 had RNA sequencing available. Sample preparation, DNA sequencing, and RNA sequencing for each assay were conducted as previously described (28–32).

Of these samples, 2,058 breast cancer and 1,216 ovarian cancer FFPE tumor samples with at least 20% tumor purity underwent tumor-normal matched DNA sequencing with the latest xT version. These breast and ovarian samples were used for training, evaluation, and exploratory analyses for HRD-DNA, and were all run on the latest assay version to ensure consistent probe design and bioinformatics pipelines. Genomic data from the xT assay was analyzed for variants, fusions, rearrangements, copy number, and loss-of-heterozygosity. For HRD-RNA, all samples with RNA sequencing data were aligned to the Ensembl hg37 transcriptome using kallisto (33,34). Transcript read counts were summated to the gene level and normalized for transcript length, GC content, and library size. Batch correction was applied for samples sequenced with different probe designs (35). All samples included in these analyses passed QC metrics, including minimum read depth, mapping rate, and duplication rate.

### HRD label annotation

Before developing the HRD-DNA and HRD-RNA models, samples were labeled as BRCA*-*biallelic, HRR-wild-type (WT), or HRD-ambiguous based on their mutational status for both *BRCA1/2* and a subset of HRR-related genes. Samples with biallelic loss of *BRCA1* or *BRCA2* were labeled as BRCA-biallelic. Biallelic loss was defined as either (a) homozygous deletion, (b) a pathogenic germline or pathogenic somatic mutation with overlapping LOH of the other allele, or (c) a co-occuring pathogenic germline and pathogenic somatic mutation. HRR-WT samples were defined as samples that had no detected pathogenic mutations, including variants with a low variant allele frequency (VAF), variants of unknown significance (VUS), fusions, copy loss, or LOH in *BRCA1, BRCA2, CDK12, PALB2, RAD51B, RAD51C*, or *RAD51D*. Samples that did not meet the criteria for the BRCA-biallelic or HRR-WT groups were labeled HRD-ambiguous, which fell into two major categories: *BRCA1/2* monoallelic loss or HRR mutated with any alteration in *CDK12, PALB2, RAD51B, RAD51C*, or *RAD51D*. Samples with mutations in these HRR genes were excluded from the HRR-WT group based on their reported associations with HRD status and enrichment for HRD+ calls in initial model iterations (36–42). Samples from patients treated with PARP inhibitors at any point in their clinical history were also considered HRD-ambiguous, regardless of mutation status. Additionally, samples with BRCA reversion mutations (identified by clinical scientists) were also considered HRD-ambiguous. These HRD-ambiguous samples were excluded from model training, development, and evaluation, but were used for exploratory analyses. Overall, ∼75% of eligible samples were considered HRD-ambiguous (Figure 1, Supplemental Figure 1).

**Figure 1.**
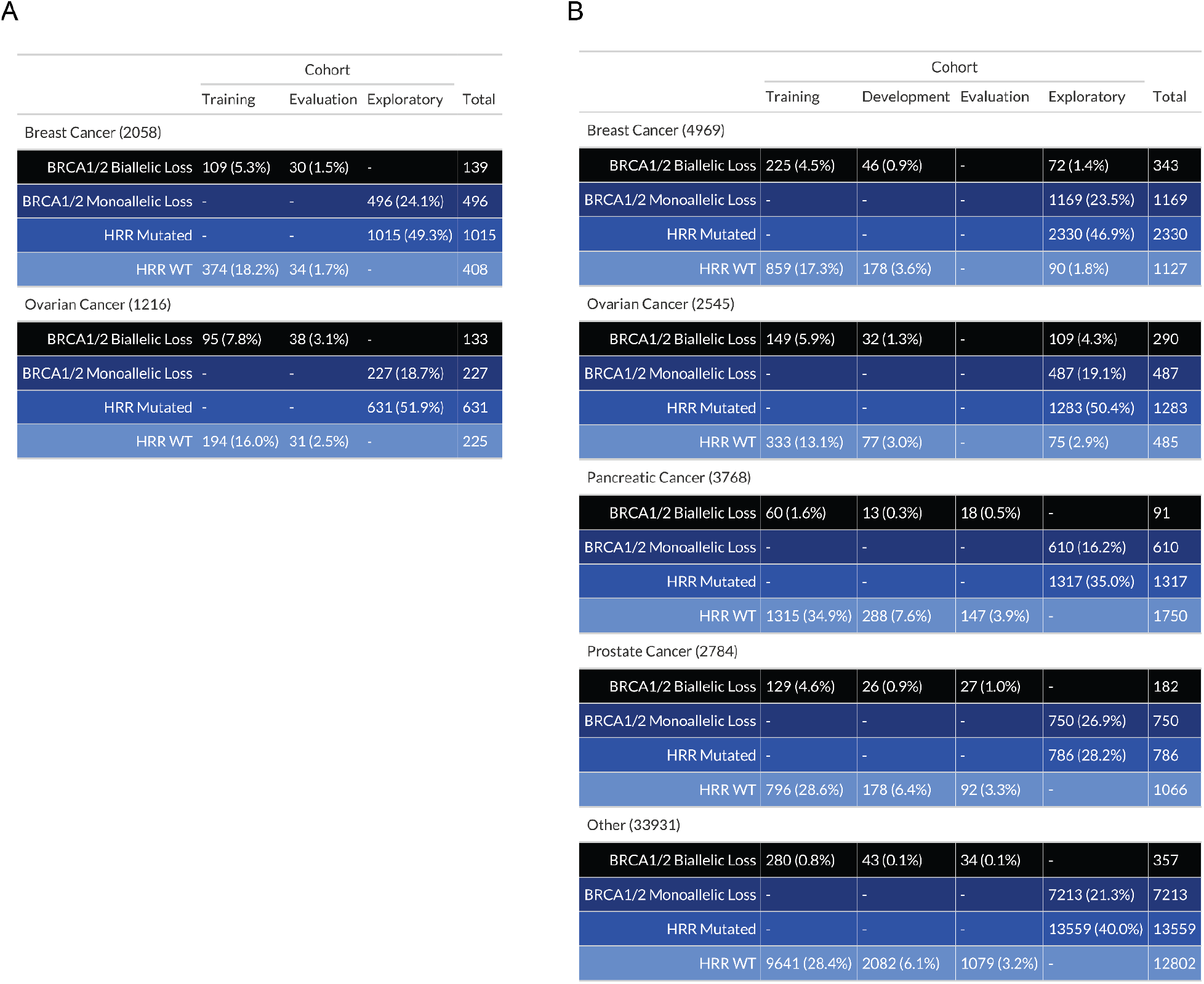
Sample composition for model training, development, evaluation, and exploratory sets by HRR mutation status and cancer type. (A) **HRD-DNA**. (B) **HRD-RNA**. To train, develop and evaluate the HRD-DNA and HRD-RNA models, *BRCA1/2* biallelic loss and HRR-WT samples were randomized into the training, development and evaluation sets, while samples with *BRCA1/2* monoallelic loss or HRR alterations (monoallelic or biallelic) in a select number of genes, were assigned to the exploratory set. Samples in the evaluation set were used to test final model performance. Samples in the exploratory set were used to determine overall rates of samples called HRD+ and enrichment of HRR mutations in HRD+ calls. Development sets were only utilized in the HRD-RNA test. Percentages were calculated as a function of the total samples in each cancer cohort for each test.

### DNA variant, copy number, loss-of-heterozygosity, and fusion annotations

DNA variants, copy number, loss-of-heterozygosity, and fusions were annotated using a combination of bioinformatics pipelines and manual clinical scientist filtering, as described in Beaubier *et al*. (28). A minimum 5% variant allele frequency (VAF) was used for calling variants. All BRCA reversion mutations were identified by clinical scientists. Samples that had variants with <5% VAF in select homologous recombination genes of interest were considered to be HRD-ambiguous. To determine homozygous deletions, samples required either (a) evidence of four consecutive probe regions or (b) 20% of the probed length of the gene to have evidence of deletion. A germline or somatic variant with LOH was considered as a biallelic loss when LOH was detected in the same probe region as the detected variant. For fusion events to pass quality control, the fusion was required to present at least five reads of evidence within the DNA-seq data.

### Calculation of gwLOH

The gwLOH calculation required calculation of aneuploid regions followed by the calculation of the total fraction of bases within probe regions with observed LOH. Percent probe loss was calculated as the number of probes with evidence of LOH on a chromosome arm divided by the total number of probes for the chromosome arm. After excluding probe regions on chromosome arms with ≥80% probe loss or sex chromosomes, gwLOH was calculated as the total number of sequenced bases in probe regions with LOH divided by total number of bases covered by all probe regions within the assay. Regions with homozygous deletions were considered to have LOH.

### HRD-RNA model preprocessing and training

47,997 samples were eligible for HRD-RNA model development; BRCA-biallelic and HRR-WT samples were stratified by cancer type and HRR mutation status and then randomly assigned at a 12:2:1 ratio to the training, development, and evaluation sets, respectively (Figure 1B). Normalized gene abundance values for each gene were standardized by removing the mean and scaling to unit variance. These gene expression values were the input to a logistic regression model with L2 regularization and weighting of the positive class. The optimal regularization strength, positive class weighting, and number of genes was determined through repeated training and evaluation using the training and development sets. Each repetition used eleven of the twelve training folds to learn the mean and variance scaling parameters and train the model. This preprocessing and modeling pipeline was evaluated on the development set. Each of the twelve folds in the training data was excluded exactly once. After all twelve repetitions, the F1-scores were averaged together to create a single score for a single set of hyperparameter values, which served as the objective function for hyperparameter tuning. 50 sets of hyperparameters were evaluated; five were randomly seeded to ensure sufficient coverage of the hyperparameter space. During hyperparameter tuning, the next set of hyperparameter values to evaluate was selected using Bayesian optimization based on the mean F1-scores of the preceding hyperparameter evaluations (43). Tuning the HRD-RNA model revealed the optimal set of hyperparameters as an inverse regularization strength of 0.0009 (strong penalty for large parameters), class weight of 11.182 (upweighting of the HRD+ class), and 20,000 genes (Supplemental Figure 7). These hyperparameter values were used to train the final HRD-RNA model on all twelve training folds. The HRD-RNA model was implemented in python using the *sklearn* logistic regression function with default parameters except as specified above (44).

### Transformation into HRD-RNA Scores

The final HRD-RNA score was created by transforming the HRD-RNA logistic regression log-odds values using a logistic function with a maximum value of 100, a logistic growth rate of 1, and a midpoint of 0.72. The midpoint was chosen to optimize the F1-score for distinguishing BRCA-biallelic and HRR-WT samples on the combined training and development sets (Figure 3A). The final HRD-RNA scores have values from 0 to 100, with a score of less than 50 indicating a prediction of HRD-, and a score of greater than or equal to 50 indicating a prediction of HRD+.

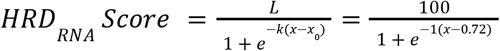

**Figure 2.**
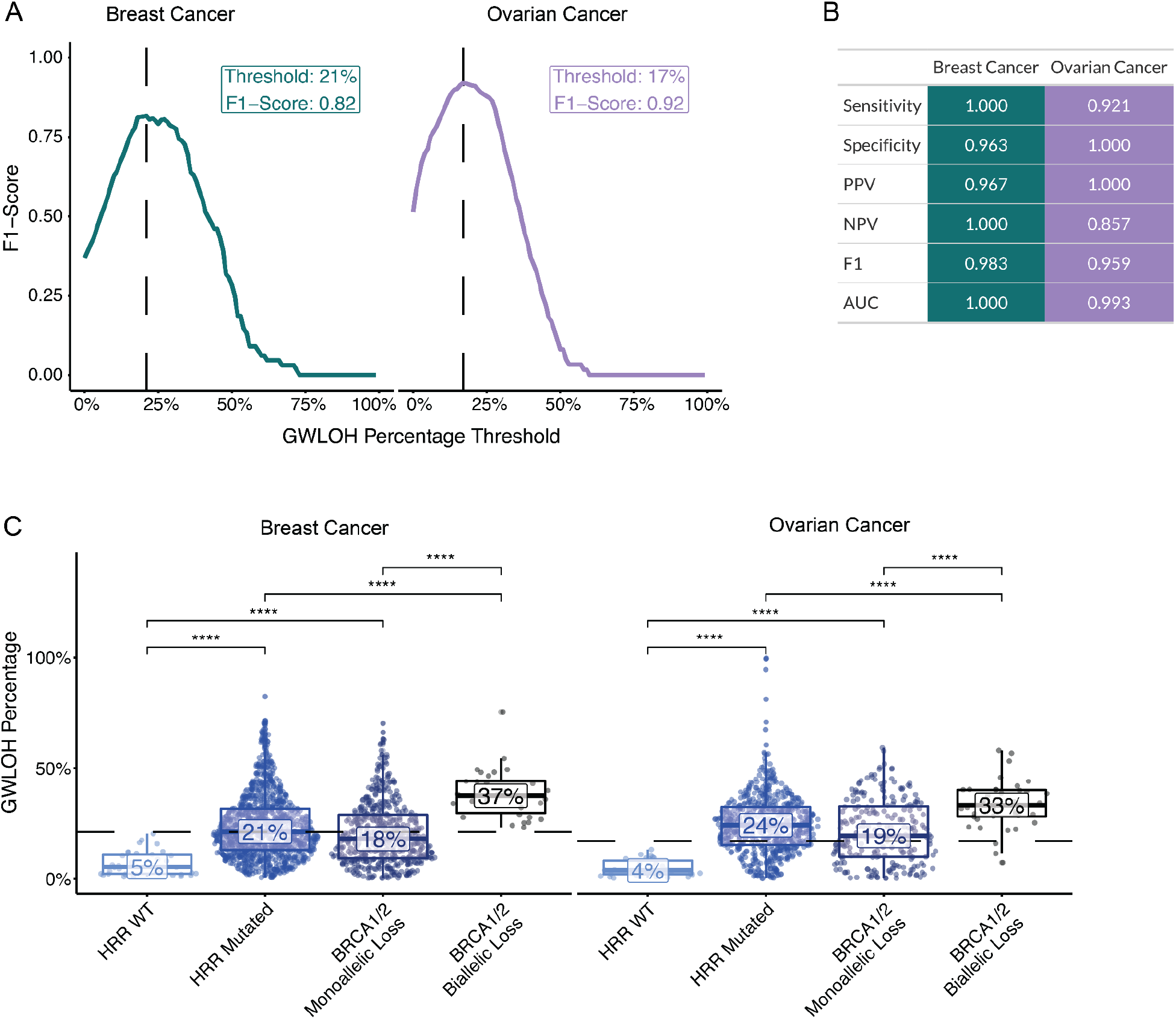
The HRD-DNA model predicts HRD status from gwLOH. (A) Thresholds for calling a sample HRD+ for breast and ovarian cancer were set based on the maximum F1-score within samples in the training set. (B) Metrics used to assess HRD-DNA performance within the evaluation set. (C) Distribution of gwLOH scores across different HRR genotypes in the evaluation cohort. HRR mutated samples contain samples with either monoallelic or biallelic loss in a select number of HRR genes. Values in the box represent the median gwLOH percentage within each HRR genotype (**** p-value < 0.0001 for Wilcoxon test). Dotted lines are the thresholds chosen in (A) for each cancer type. Statistical differences between HRR-WT and BRCA biallelic loss were not shown, but all were significant (p-value < 0.0001 for Wilcoxon test).

**Figure 3.**
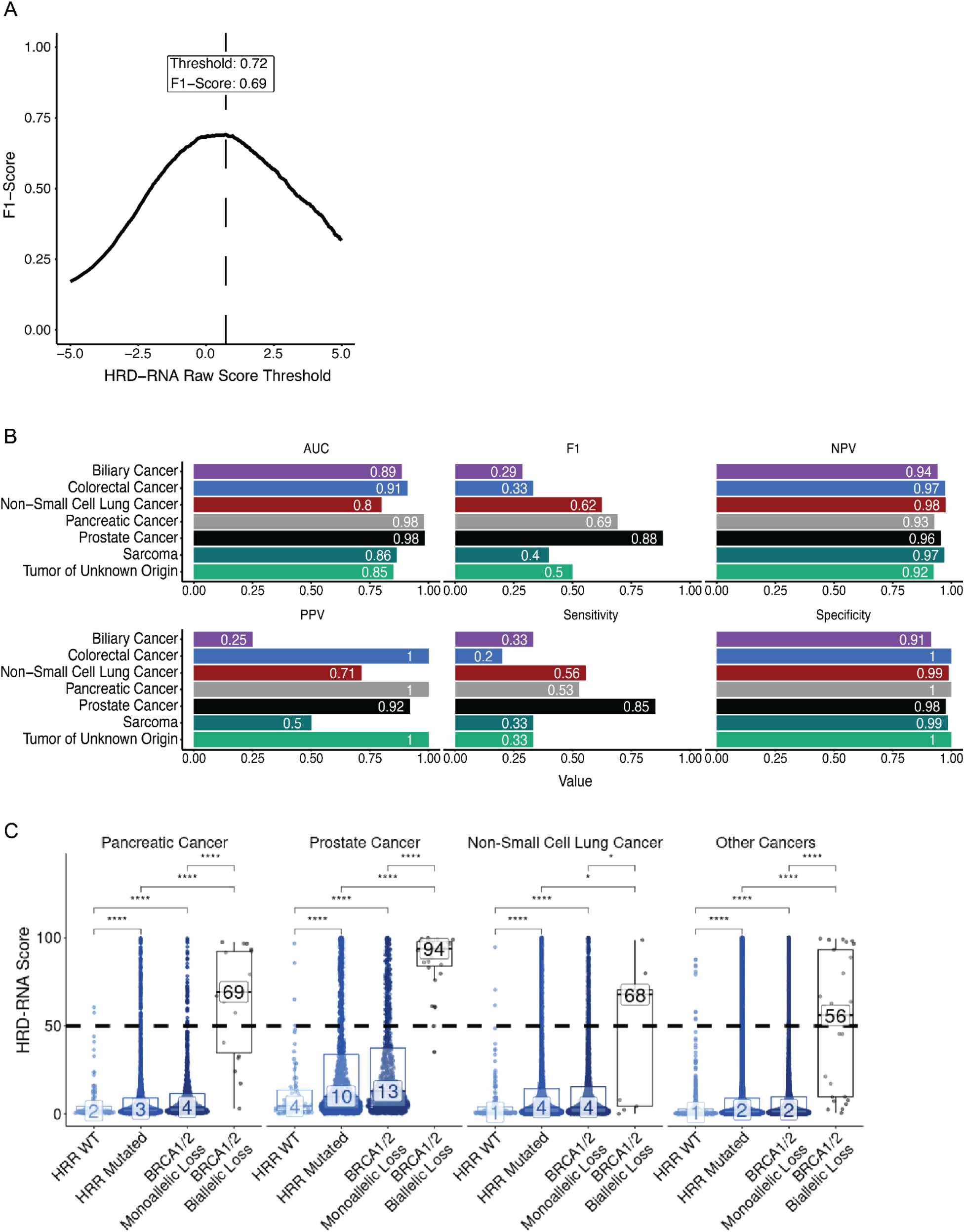
The HRD-RNA model determines HRD-status for cancer cohorts outside breast and ovarian cancer using a logistic regression model trained on RNA-seq data. (A) The threshold for calling a sample HRD+ was set as the raw RNA score that had the maximum F1 score on the training and development samples. For final reporting, the raw score was transformed to the final HRD-RNA, where a score of 50 represents the chosen threshold (Methods). (B) Metrics used to assess HRD-RNA model performance across cancer cohorts for samples within the evaluation cohort. (C) Distribution of HRD-RNA scores across different HRR genotypes in the evaluation cohort for cancer indications with > 3 BRCA-deficient samples in the evaluation cohort. Dotted line represents the threshold chosen in (A). Values in the box represent the median HRD-RNA score. Differences determined by Wilcoxon test (* p-value < 0.05, **** p-value < 0.0001). Statistical differences between HRR-WT and BRCA biallelic loss were not shown. Statistical differences between HRR-WT and BRCA biallelic loss were not shown, but all were significant (p-value < 0.0001 for Wilcoxon test). AUC: Area Under the Curve. NPV: Negative Predictive Value.

### Statistical analyses

One-way Fisher’s exact tests were used to test for enrichment, i.e. enrichment of BRCA-monoallelic samples with HRD+ calls. To correct multiple hypothesis testing, Bonferroni correction was used to calculate a false-discovery rate, implemented by the *p*.*adjust()* function from the *stats* package in R (45,46). All correlation coefficients and p-values represent Pearson correlations implemented by the *cor*.*test()* function from the *stats* package (46).

### Data Availability Statement

Raw data for this study were generated at Tempus Labs. Derived data supporting the findings of this study are available within the paper and its Supplementary Figures or available from the authors upon request.

## Results

### HRD-labels assigned based on HRR-gene mutation status

A superset of 48,843 tumor samples with targeted DNA-seq and full-transcriptome RNA-seq was used in either model training, development, evaluation, or exploration for the HRD-DNA or HRD-RNA models (Figure 1). Samples were labeled as either HRD+, HRD-ambiguous, or HRD-, with HRD+ defined as BRCA-biallelic, HRD-as no mutations in *BRCA1, BRCA2, CDK12, PALB2, RAD51C*, or *RAD51D*, and HRD-ambiguous as BRCA-monoallelic or HRR-mutated (Methods). For reference, samples with any mutation or LOH in were considered to be HRD-ambiguous and were not included in the model training, development, or evaluation sets. HRD+ and HRD-labeled samples were randomized into the training, development, and evaluation sample sets, while HRD-ambiguous samples were considered exploratory.

Among breast cancer samples, 6.8% were annotated as BRCA-biallelic for both the HRD-DNA and HRD-RNA models, respectively. Additionally, 10.9% and 11.5% of ovarian cancers were annotated as BRCA-biallelic for the HRD-DNA and HRD-RNA model cohorts, respectively (Figure 1A). These prevalences are slightly lower than what has previously been observed for breast (7.8%) and ovarian (20%) cancers (47–49). This discrepancy may be attributable to our requirement for biallelic BRCA alterations and cohort differences found in the Tempus Oncology Database (49,50). Our observed prevalence of *BRCA*-deficiency in prostate and pancreatic cancers is 6.5% and 2.4%, which is similar to the values reported by others: 6.2% and 3.4% respectively (51,52).

For the approximately 75% of samples that were designated as HRD-ambiguous, the majority of samples were excluded from the HRR-WT class due to monoallelic LOH in either *BRCA1/2* or an HRR gene (Supplemental Figure 1). Samples with LOH were included in the HRD-ambiguous class given the potential for co-occurring alternative mechanisms of BRCA-deficiency (*i*.*e*., promoter methylation, a low variant allele frequency [VAF], or a variant of unknown significance [VUS]), limiting the ability to apply a high-confidence HRD label.

### Aneuploidy exclusion and gwLOH calculations for HRD-DNA

The HRD-DNA model was designed to predict the HRD status of breast and ovarian tumor samples using gwLOH, excluding aneuploid chromosome arms. To detect aneuploidy and correct for this potential confounder, we determined the optimal probe loss fraction associated with chromosome arm deletion (and therefore excluded from the gwLOH calculation) and the optimal gwLOH threshold to distinguish BRCA-biallelic (HRD+) samples from HRR-WT (HRD-) samples for both breast and ovarian cancers (Supplemental Figure 2A). We found that model performance, measured by F1-score (the harmonic mean of precision and recall), was more sensitive to the gwLOH threshold than probe loss threshold, and optimal probe loss thresholds were 78% and 84% for breast cancer and ovarian cancer, respectively. For biological consistency, and given the breast cohort is approximately twice as large as the ovarian cohort, a probe loss threshold of 80% was selected to identify chromosome arms lost due to aneuploidy for both cohorts. The chosen probe loss threshold was validated by applying it to glioma samples that received fluorescence *in situ* hybridization (FISH) to assess genetic deletion of chromosome 1p (Supplemental Figure 2B), which is often used to diagnose oligodendrogliomas (53–55). Samples with negative 1p FISH results had a significantly lower fraction of probes lost compared to 1p FISH positive (p < 2.2e-16) (Supplemental Figure 2C). The chosen probe loss threshold of 80% achieved 89% concordance with 1p FISH results, and samples with >80% probe loss were significantly enriched for 1p FISH positivity (Fisher’s exact test, p-value = 1e-31) (Supplemental Figure 2D). Together, these results demonstrate that the probe loss method accurately identifies chromosome arms lost due to aneuploidy.

Finally, the training samples were used to identify the optimal gwLOH score—excluding aneuploid chromosome arms—for calling a sample HRD+ or HRD-. The gwLOH threshold was determined as the threshold that best distinguished the BRCA-biallelic from HRR-WT samples, measured by F1-score. The optimal gwLOH threshold was 21% and 17% for breast and ovarian cancer, respectively (Figure 2A). The evaluation samples were then used to evaluate the performance of the chosen probe loss and gwLOH thresholds, yielding robust performance metrics: sensitivity (breast = 1, ovarian = 0.921), specificity (breast = 0.963, ovarian = 1.0), positive predictive value (PPV) (breast = 0.967, ovarian = 1.0), negative predictive value (NPV) (breast = 1.0, ovarian = 0.857), F1-score (F1: breast = 0.983, ovarian = 0.959), and AUC (breast = 1.0, ovarian = 0.993) (Figure 2B). The HRD-DNA model had a lower sensitivity and NPV for ovarian cancer relative to breast cancer, suggesting there may be a greater fraction of patients with low gwLOH that are HRD+ in ovarian cancer (Figure 2C). Finally, HRR-WT samples had significantly lower gwLOH compared to HRD-ambiguous samples in both HRR-mutated (Wilcoxon test; p-value_breast_ = 2e-14, p-value_ovarian_ = 4e-16) and *BRCA1/2* monoallelic samples (Wilcoxon test; p-value_breast_ = 1e-10, p-value_ovarian_ = 4e-11). Samples were predicted HRD+ at a lower rate for HRR-WT samples (breast = 2.9%, ovarian = 0%) compared to HRR-mutated (breast = 56.7%, ovarian = 69.3%) and *BRCA1/2* monoallelic (breast = 48.8%, ovarian = 56.8%) — suggesting other potential drivers of the HRD-phenotype (Supplemental Figure 3A). The gwLOH biomarker accurately separated samples with *BRCA1/2* biallelic loss from samples with no evidence of mutations in *BRCA1/2* or a subset of HRR genes in both breast and ovarian cancer.

### HRD-RNA model training and evaluation

BRCA-biallelic and HRR-WT samples from all cancer types were included in the training and development sets for the HRD-RNA model, including breast and ovarian cancers (Figure 1B). The HRD-RNA model was evaluated for cancer types that included at least 3 BRCA-biallelic samples in the evaluation set (Figure 3B). Across these cancer types, the model achieved a PPV of 25%, indicating that only a fraction of the patients predicted HRD+ exhibited *BRCA1/2*-deficiency. The highest AUCs on the evaluation set were in prostate (0.98) and pancreatic (0.98) cancer, which is unsurprising given that tumor pathogenesis in these cohorts has been previously associated with BRCA status (56,57). For prostate and pancreatic cancers in the evaluation and exploratory sets, there was strong separation between the BRCA-biallelic and HRR-WT samples (Wilcoxon test; p-value_Prostate_ = 1e-14, p-value_Pancreatic_ = 6e-11), and between the HRR-WT and HRD-ambiguous samples (Wilcoxon test; p-value_Prostate_ = 4e-9, p-value_Pancreatic_ = 6e-8) (Figure 3C).

While other cancer cohorts — biliary, colorectal, NSCLC, sarcoma, and cancers of unknown primary (tumors of unknown origin) — had a lower prevalence of *BRCA1/2* alterations, previous work has suggested that these cohorts may exhibit the HRD phenotype and respond to PARP inhibitors (Figure 1B) (58–60). We hypothesized that tumors from patients with these cancer types may exhibit the HRD phenotype in the absence of BRCA loss. For these cohorts, we observed lower overall performance relative to BRCA status when compared to pancreatic and prostate cancer (Figure 3B).

Across all cohorts evaluated using the HRD-RNA model, the number of samples with BRCA-biallelic loss in the training and development sets was positively correlated with the F1-score (R = 0.92, p-value = 3e-3) and sensitivity (R = 0.95, p-value = 1e-3) of the model on the evaluation set (Supplemental Figure 4). This result suggests a biological and/or modeling constraint in cancer cohorts with few BRCA-biallelic samples — either BRCA deficiency may be a poor surrogate for HRD status or there may be insufficient BRCA-biallelic samples to identify a signal. The high fraction of samples that were HRD-ambiguous and predicted HRD+, i.e. 13.1% of BRCA-monoallelic and 11.7% of HRR-mutated prostate cancers, suggests that mutations in other HRR pathway genes or epigenetic modifications (*i*.*e*., hypermethylation) may drive the HRD-phenotype in cancer types not traditionally associated with BRCA-status (Supplemental Figure 3; Figure 3C).

### The HRD-DNA and HRD-RNA models capture an underlying HRD-phenotype consistent with the literature

Both the HRD-DNA model and the HRD-RNA model captured biallelic loss of *BRCA1/2* with high sensitivity and specificity in BRCA-associated tumors (breast, ovarian, pancreatic, and prostate cancer) (Figure 2B; Figure 3B). We hypothesized that some fraction of the ∼75% of samples annotated as HRD-ambiguous would be predicted HRD+ (Figure 1; Supplemental Figure 1); these HRD-ambiguous samples had higher HRD scores (Figure 2C, Figure 3C) and a higher frequency of predicted HRD+ compared to HRR-WT samples (Supplemental Figure 3) by both the HRD-DNA and HRD-RNA models. Together, these findings further suggest that there is an underlying HRD-phenotype not captured by BRCA biallelic loss alone.

Outside *BRCA1/2* alterations, samples predicted HRD+ by each HRD model were enriched for biallelic loss of HRR genes, which would suggest an alternative mechanism for the HRD-phenotype (Figure 4A). Significant enrichment of HRD-DNA+ predictions was observed in samples with biallelic loss in *ATM* (FDR = 2e-11), *ATRX* (FDR = 3e-3), *BARD1* (FDR = 1e-3), *BRIP1* (FDR = 4e-13), *CDK12* (FDR = 2e-6), *FANCA* (FDR = 8e-3), *MRE11* (FDR = 9e-4), *PALB2* (FDR = 3e-6), or *RAD51D* (FDR = 2e-8). Previous studies have also demonstrated that biallelic loss in *BARD1, BRIP1, FANCA, MRE11, PALB2*, and *RAD51D* is associated with a higher gwLOH score in BRCA-associated cancers (9). In other cancer types, there was a lower overall fraction of samples predicted HRD+ across HRR genes, highlighting the lower frequency of HRD in these cancer types (Figure 4A). However, there was significant enrichment for HRD-RNA+ predictions in samples with biallelic loss of *ATRX* (FDR = 9e-8), *CDK12* (FDR = 4e-5), *FANCA* (FDR = 2e-2), *PALB2* (FDR = 3e-8), and *RAD51B* (FDR = 5e-4). Enrichment of HRD+ calls in samples with biallelic loss of HRR genes highlights the utility of both the HRD-DNA and HRD-RNA models in identifying a number of potential drivers of the HRD phenotype that are independent of *BRCA*1/2 biallelic loss.

**Figure 4.**
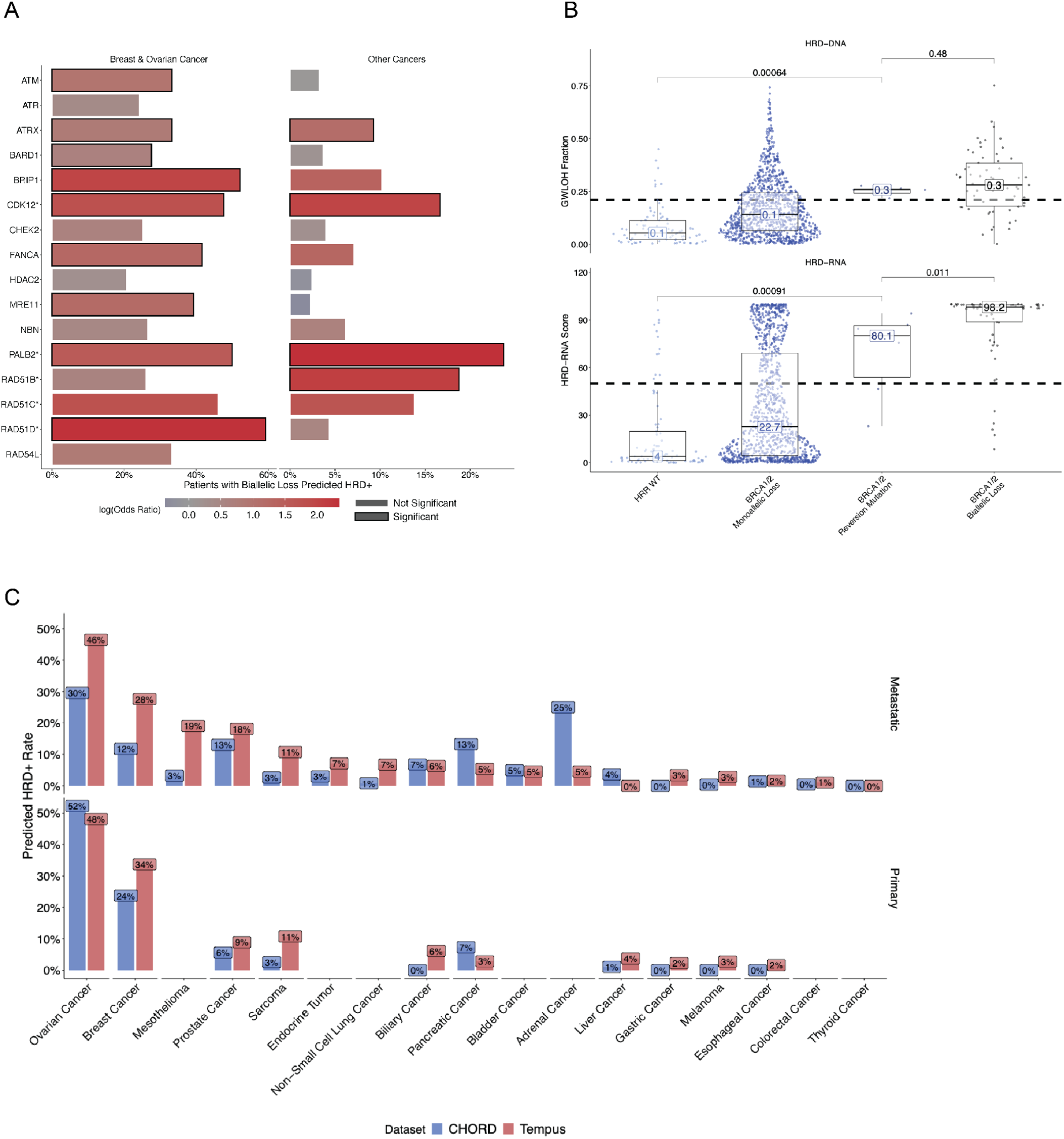
The HRD-DNA and HRD-RNA models are enriched for a HRD-phenotype and are concordant with published HRD+ rates. (A) Enrichment for HRD+ calls in samples with biallelic loss of HRR genes for breast cancer and ovarian cancer (HRD-DNA), and other cancers (HRD-RNA). Enrichment was calculated using a Fisher’s exact test comparing samples from the HRD-ambiguous and HR-WT samples that has biallelic loss of specific HRR gene versus all other samples. Significance was determined as p-value < 0.05. (B) Distribution of GWLOH percentage (top) and HRD-RNA scores (bottom) of breast cancer samples with *BRCA1/2* reversion mutations. Significance shown for two-sided Wilcoxon-test. (C) Predicted rates of HRD+ samples across cancer types compared to published rates (CHORD), stratified by primary and metastatic samples.

**Figure 5.**
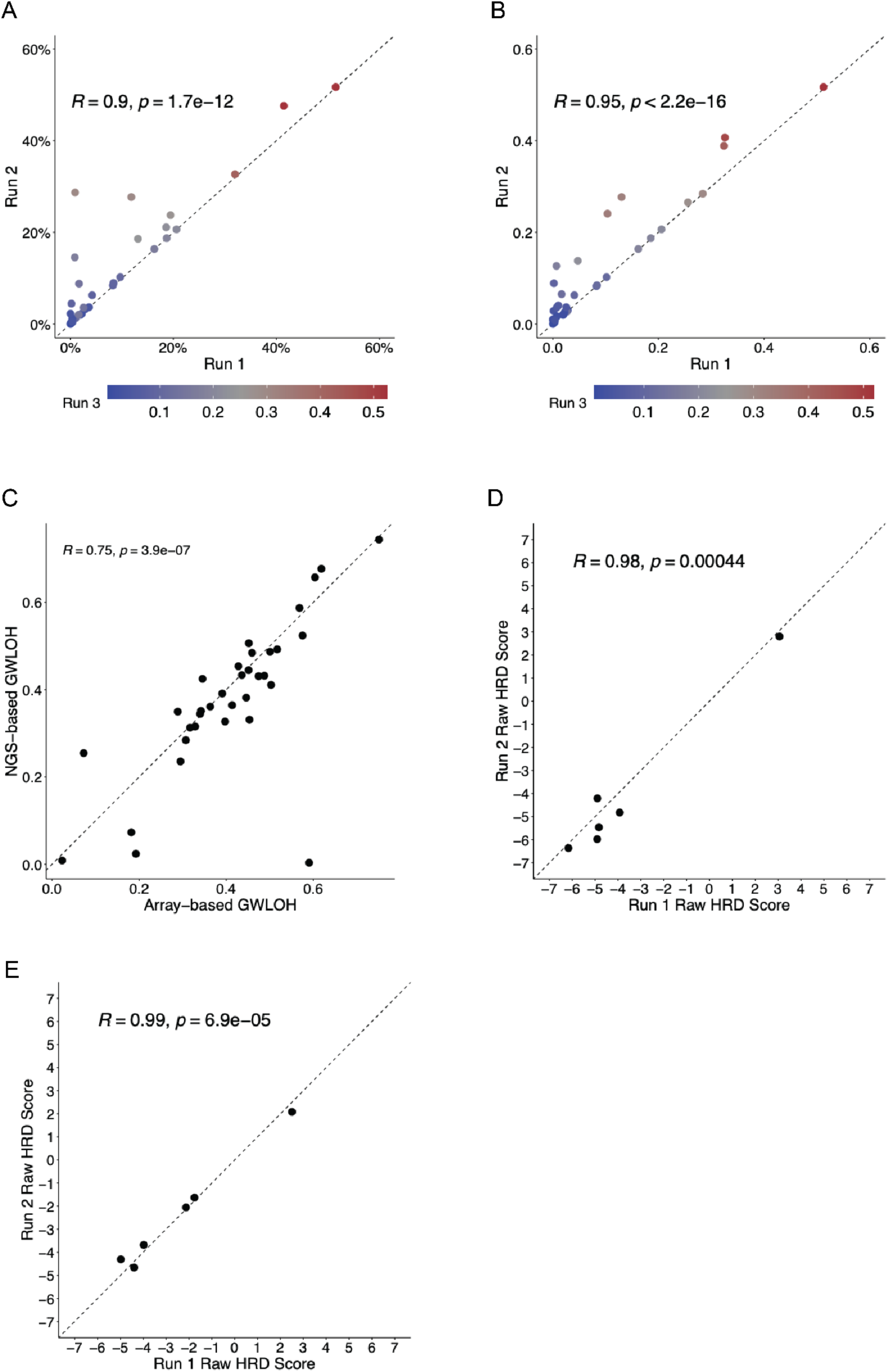
Inter- and intra-assay concordance for the HRD-DNA and HRD-RNA models. (A) Intra-assay concordance of HRD-DNA for 32 samples each run in triplicate. (B) Inter-assay concordance of HRD-DNA for 34 samples each run in triplicate. (C) Correlation between HRD-DNA and Omni 2.5 BeadChip array based genome-wide LOH. (D) Intra assay concordance of HRD-RNA for 6 samples each run in duplicate. (E) Inter assay concordance of HRD-RNA for 6 samples each run in duplicate. Dotted lines are identity lines.

The HRD-RNA model, in contrast to the HRD-DNA model, has the potential to capture a dynamic HRD phenotype. For example, one mechanism for PARP inhibitor resistance is via a *BRCA1/2* reversion mutation (61–63). Though rare, samples with these mutations can serve as a test of the dynamic nature of the HRD-RNA model compared to the HRD-DNA model. In the presented data, breast cancer samples most frequently possessed BRCA reversion mutations (n = 7); these cases were excluded from the HRD-RNA model training and development sets. The HRD-RNA model predicted lower HRD scores for samples with a *BRCA1/2* reversion mutation compared to samples with biallelic loss of *BRCA1/2* (Wilcoxon test; p-value = 0.002), indicating the ability of the HRD-RNA model to capture a dynamic HRD phenotype (Figure 4C). On the other hand, there was no significant difference in the HRD-DNA score between samples with a *BRCA1/2* reversion mutation and BRCA-biallelic samples. Although five out of the seven samples with BRCA reversions were predicted to be HRD+ by HRD-RNA, this may be attributable to clonal BRCA reversion (mean VAF: 13.3%), resulting in clonal HRD.

Finally, given the different approaches for predicting HRD status (HRD-DNA and HRD-RNA) and the low PPV for non-BRCA associated cancer cohorts, we compared rates of HRD+ calls between the presented models and the literature (Figure 3B, Figure 4B). The Classifier of HOmologous Recombination Deficiency (CHORD) method utilizes genomic features from whole-genome sequencing data with a random forest classifier to determine HRD-status (8). HRD prevalence across tumor types estimated by CHORD was used as a benchmark for positivity rates predicted by HRD-DNA (breast and ovarian) and HRD-RNA (all other cohorts), stratified by primary and metastatic status. For both primary and metastatic cancers, there was a strong, positive correlation (HRD-DNA: R^2^ = 0.63, p-value = 4e-4; HRD-RNA: R^2^ = 0.83, p-value = 1e-7) between the predicted frequency of HRD+ samples in the presented models and the respective predicted frequency in CHORD (Figure 4C; Supplemental Figure 5). This suggests predicted HRD+ prevalence within each cancer type is supported by other studies.

There were a few notable deviations in predicted HRD prevalence across tumor types. First, the HRD-DNA model reported an HRD+ prevalence of 52% and 61% in primary breast and ovarian cancer, while CHORD presented 54% and 30%, respectively. This difference may be due to the fact that patients sequenced by Tempus are often later-stage and have received more lines of therapy. Moreover, other groups have reported ovarian cancer HRD+ prevalence in the 40-50% range (64,65). In metastatic breast cancer, CHORD reported an HRD+ prevalence of 12%, while the HRD-DNA model predicted 46%. Upon further inspection of the cohort used by CHORD, 13.5% of all metastatic breast cancer samples were triple-negative (TNBC), the breast subtype for which HRD positivity is highest (66). By contrast, the Tempus cohort is enriched for TNBC samples given that these patients have few treatment options and worse outcomes, and are thus more likely to undergo Tempus NGS testing. Indeed, nearly 24% of Tempus breast cancers are TNBC, and, as a result, models predicting HRD status using Tempus data should be expected to report a higher HRD+ prevalence compared to CHORD.

### HRD-DNA and HRD-RNA are robust to confounders

The limit of detection (LOD) for both the HRD-DNA and HRD-RNA models was determined by calculating the sensitivity of both models across different tumor purities. The LOD was set as the lowest tumor purity at which the positive predictive value (PPV) was greater than 70%. For the HRD-DNA model, PPV was calculated using all breast or ovarian samples that had either *BRCA1/2* biallelic loss or were BRCA-WT. A tumor purity threshold was set at 40% based on the PPV threshold (Supplemental Figure 6). For the HRD-RNA model, PPV was calculated using all samples, including breast and ovarian samples, that had either *BRCA1/2* biallelic loss or were BRCA-WT and were in the evaluation set. A tumor purity threshold was set at 30% based on the PPV threshold (Supplemental Figure 6). These data indicate that the HRD-DNA and HRD-RNA models are performant among samples with at least 40% and 30% tumor purity, respectively.

To test whether model performance is tissue-site dependent, the evaluation set was stratified by sample status - primary, metastatic, lymph node, or unknown (Supplemental Table 1). For both HRD-DNA and HRD-RNA, model performance was similar in each sample stratification. For HRD-DNA, F1-score ranged from 0.933-1.0 and sensitivity ranged from 0.875-1.0. For HRD-RNA, F1-score is higher in primary (0.741) compared to metastatic (0.591) samples, mostly driven by higher sensitivity in primary samples (0.690) compared to metastatic (0.448) samples. However, AUC was similar between primary (0.956) and metastatic (0.953) samples, compensated for the higher specificity in metastatic (0.994) samples over primary (0.990) samples. Overall, HRD-DNA and HRD-RNA performance is robust to biopsy site.

Finally, to establish reproducibility across sequencing runs, experiments were run to demonstrate inter- and intra-assay concordance for the HRD-DNA and HRD-RNA models. To demonstrate intra-assay concordance for the HRD-DNA model, 32 samples were sequenced in triplicate in the same DNA sequencing run using the same reagent lot with different barcodes (Figure 6A). There was a significant correlation for all run comparisons (.87 < R^2^ < 0.97 across all comparisons, p-value < 1e-11), demonstrating intra-assay concordance. To demonstrate inter-assay concordance for HRD-DNA, 34 samples were sequenced in triplicate on different days, using different instruments, different lab technicians, and at least two manufacturing reagent lots (Figure 6B). There was a significant correlation for all run comparisons (.89 < R^2^ < 0.98 across all comparisons, p-value < 1e-12), demonstrating high intra-assay concordance. To orthogonally validate HRD-DNA gwLOH calls, 34 samples were run via Omni2.5 BeadChip copy number array at an external lab (67). The HRD-DNA score was calculated from both sets of copy calls and was highly correlated (R^2^ = 0.75, p-value = 3.9e-7; Figure 6C).

To demonstrate intra-assay concordance for the HRD-RNA model, 6 samples were sequenced in duplicate in the same RNA sequencing run using the same reagent lot with different barcodes (Figure 6D). There was a significant correlation of the raw RNA output (R^2^ = 0.98, p-value = 4.4e-4), demonstrating intra-assay concordance. To demonstrate inter-assay concordance for the HRD-RNA model, 6 samples were sequenced in duplicate on different days, using different instruments, different lab technicians, and at least two manufacturing reagent lots (Figure 6E). There was a highly significant correlation of the raw RNA output (R^2^ = 0.99, p-value = 6.9e-5), demonstrating intra-assay concordance.

## Discussion

Here, we present two models, HRD-DNA and HRD-RNA, that predict the HRD status of clinical FFPE samples. While the models were trained and evaluated on their ability to predict *BRCA1/2*-biallelic from HRR-WT samples, HRD+ samples were also enriched for biallelic loss of other HRR genes, and predicted frequencies of HRD+ samples across cancer cohorts are largely in agreement with what has been reported in the literature. The models’ ability to detect an HRD phenotype rather than solely BRCA-loss is critical outside of the so-called non-BRCA associated tumors (breast, ovarian, pancreatic and prostate). Such HRD detection tools also present an opportunity to identify alternative drivers of the HRD phenotype, and potential indications beyond those reported in the literature. For example, we found high rates of HRD in sarcomas and mesotheliomas, where RNA-based approaches may be particularly useful. Together, these models provide new biomarkers for HRD-status in solid tumor samples.

The basis for the HRD-DNA model, gwLOH, is known to have different manifestations across tumor types, necessitating cancer-specific thresholds for calling a sample HRD+. For breast and ovarian cancers, there were sufficient BRCA-biallelic samples to set a threshold for HRD-DNA to predict HRD-status. However, other cohorts either had too few BRCA*-*biallelic samples or little difference in gwLOH between BRCA-biallelic and HRR-WT samples, necessitating an alternative approach. For solid tumor indications outside of breast and ovarian, the HRD-RNA model predicts HRD-status using a logistic regression model trained on bulk RNA-seq data from solid tumor FFPE samples. Breast and ovarian cancer had the highest prevalence of BRCA-biallelic samples (Figure 1); despite these being the dominant cancer cohorts for HRD+ samples, we assumed that the transcriptional signature of *BRCA1/2* mutations and HRD would be consistent across tumor types for predictive power of HRD independently of tumor type. The positive relationship between the number of BRCA-biallelic samples within a tumor type and the model F1-score highlights that model performance, as benchmarked by BRCA mutation status, is superior in cohorts with higher prevalence of BRCA alterations. Cancer types with lower prevalence of BRCA-biallelic samples and a low PPV suggest that biallelic BRCA-loss may not be driving tumorigenesis. While p53 loss has been observed to confound gwLOH measurements (9), there is little understanding of confounding factors for RNA-based HRD approaches. Future work should focus on uncovering alternative causes of HRD, and disambiguating the differences in performance across cancer cohorts with the potential co-occurence of other tumor drivers where biallelic loss of BRCA may be a passenger rather than driver mutation.

The HRD-DNA and HRD-RNA models not only capture BRCA-biallelic samples but also demonstrate enrichment for samples labeled as HRD-ambiguous (Figure 2C, Figure 3C) with biallelic loss of other HRR genes (Figure 4A). While *ATM* was the most commonly lost HRR gene in ovarian, breast, and other cancers, and occurred at a rate similar to *BRCA1/2*-deficiency, HRD+ calls were enriched only in the HRD-DNA model, suggesting that *ATM* loss may be uniquely associated with a high gwLOH phenotype in breast and ovarian cancer but not HRD in other cancer types. In metastatic castration-resistant prostate cancer, clinical trials have shown little to no clinical benefit of PARP inhibitors in *ATM*-mutated patients over standard of care treatment (5,68). Other mutations were uniquely enriched in HRD+ predicted cases of breast and ovarian cancers (*BARD1, BRIP1, MRE11, RAD51D*) and other indications (*RAD51B*), suggesting additional unique drivers of HRD that may be cancer-cohort specific. Monoallelic copy loss of HRR genes was highly prevalent and could not be included in the HRD-ambiguous category without compromising the statistical power of the HRR-WT cohort. Future work should explore the association between specific alterations (i.e. SNPs, LOH, deletions), pathogenicity (VUS), contexts (i.e. germline, somatic), and genes (i.e. *BRCA1/2*, HRR genes) with HRD calls to better understand mechanisms driving the HRD+ predictions.

In both the HRD-DNA and HRD-RNA models, samples with biallelic loss of *ATRX, CDK12, FANCA*, and *PALB2* were enriched for HRD+ predictions across tumor types. Both *FANCA* and *PALB2* have been associated with increased gwLOH scores (9). Mutations in *CDK12* have been shown to confer sensitivity to PARP inhibitors in breast and ovarian cell lines (36) and clinical trials in prostate cancer (5). *PALB2* has been recognized to play a role in HRR through interactions with *BRCA1/2* (40) and, more recently, *PALB2* has been implicated as another potential genomic biomarker for HRD (39,69,70). Biallelic loss of *ATRX* has been associated with increased gwLOH in breast but not ovarian or other cancer cohorts (9). *ATRX* mutations have been shown to inhibit homologous recombination repair in cell lines (71–73), are linked to PARP inhibitor sensitivity in patient-derived xenografts (74), are associated with higher *PARP1* expression in clinical glioblastoma tumors (75), and have shown sensitivity to DNA-damaging treatment in pediatric high-grade glioma patients (76). Overall, these pan-cancer models of HRD highlight the ability to accurately capture the HRD phenotype and thus generate new hypotheses for other potential genetic drivers of HRD.

Notably, the HRD-RNA model predicted samples with BRCA-reversions to have a significantly lower HRD-RNA score than BRCA-biallelic samples, which was not true of HRD-DNA. This result suggests an RNA-based measure of HRD may capture dynamic changes in HRD phenotype upon tumor evolution. Given that PARP inhibitor resistance can be caused by a number of mechanisms that cannot be detected by DNA alone (77), future work should explore the utility of RNA-based approaches for both identifying HRD samples and tracking the emergence of resistance.

Whole-genome sequencing (WGS) is challenging and expensive to implement in real-world clinical practice, but WGS-based models have emerged as a potentially comprehensive tool to capture HRD. We demonstrated a strong correlation between the predicted frequency of one such WGS-based method, CHORD, and HRD+ frequencies from the presented models in both primary and metastatic samples. Deviations (*i*.*e*., ovarian and breast cancer) can be partially explained by differences in HRD rates among different subtypes and enrichment of subtypes with higher rates of HRD+ samples in the Tempus Oncology Database. One notable deviation from the predicted rates of HRD+ samples is in pancreatic cancer where, for both metastatic and primary samples, the predicted rates for HRD-RNA are lower than CHORD. Reported rates of HRD in pancreatic cancer vary between 3 and 30% (27,78), where RNA-based approaches have been shown to be prognostic and identify other genetic drivers of HRD (27). Given clinical trials have demonstrated no survival benefit using olaparib to treat pancreatic cancer patients with germline BRCA alterations, there remains a poor understanding of HRD manifestation in pancreatic cancer (79).

Significantly higher rates of HRD+ in mesothelioma (metastatic) and sarcoma (primary and metastatic) were seen with the presented models compared to CHORD. PARP inhibitors are currently being explored in combination with immune checkpoint inhibitors in mesothelioma (80), where improved response was observed in patients with germline mutations in HRR genes (81). However, little work has been done exploring the role of HRD as a biomarker in sarcomas. A recent study demonstrated improved response to PARP inhibitors in patient-derived soft-tissue xenografts with high *PARP1* expression (82). Collectively these observations may suggest that HRD-RNA captures a unique HRD signature in tumors that cannot be captured by genomic sequencing. Both mesothelioma and sarcoma present new cohorts to explore HRD as a potential biomarker, but more data would be needed to determine the role of HRD and its value as a biomarker in these indications.

HRD-DNA and HRD-RNA are highly performant in differentiating BRCA-biallelic from HRR-WT samples and enriching for other genomic events in the HRR pathway. HRD-DNA and HRD-RNA together suggest that while biallelic loss of *BRCA1/2* may be sufficient to detect the majority of cases of the HRD-phenotype in breast and ovarian cancer, more work is needed to identify the relationship between genotype and HRD in other cohorts. HRD-RNA has the potential ability to disambiguate this relationship by determining a pan-cancer signature of HRD that can be applied to cancer cohorts. This may be most valuable as a precision medicine tool in tumor indications with a lower frequency of biallelic BRCA loss. Further work is warranted to determine potential HRD driver events and clinical implications of HRD status across cancer indications. Importantly, future prospective clinical studies are required to assess the clinical utility of the presented HRD biomarkers for predicting response to DNA-damage targeting therapies.

## Supporting information

Combined Supplemental Information

## Acknowledgements

The authors acknowledge valuable support and feedback from numerous individuals at Tempus Labs who have contributed to and made this work possible, including (but not limited to): Alexandria M. Bobé, Matthew Kase, Adam J. Hockenberry, and Justin Guinney.

