## Supplementary material for "Validation of Genomic and Transcriptomic Models of Homologous Recombination Deficiency in a Real-World Pan-Cancer Cohort": Combined Supplemental Information

Affiliation for all authors: Tempus Labs, Chicago, IL

Running Title: Pan-cancer HRD Model

Conflict of Interest Statement: All authors are employees of Tempus Labs.

### Supplemental figure/table captions

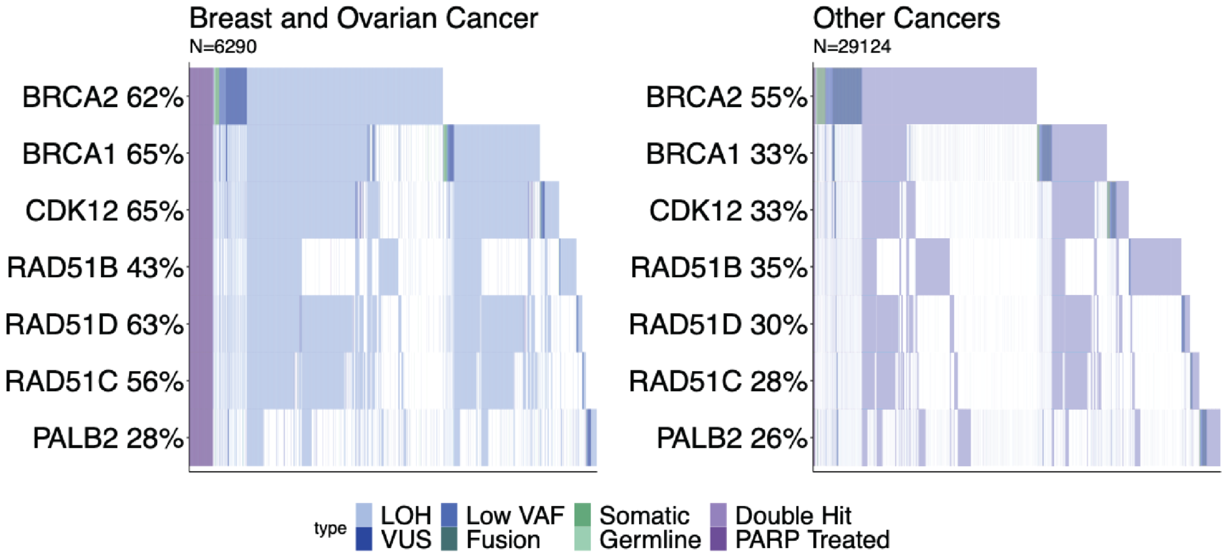


**Supplemental Figure 1. Breakdown of samples in the HRD-ambiguous sample set.** Samples were assigned to the HRD-ambiguous class based on either *BRCA1/2* monoallelic loss or mutations in a select number of HRR genes (*CDK12*, *RAD51B*, *RAD51C*, *RAD51D*, *PALB*2). Samples that had been treated with a PARP inhibitor either prior to or after sequencing were also included in the HRD-ambiguous class (labeled here PARP treated).

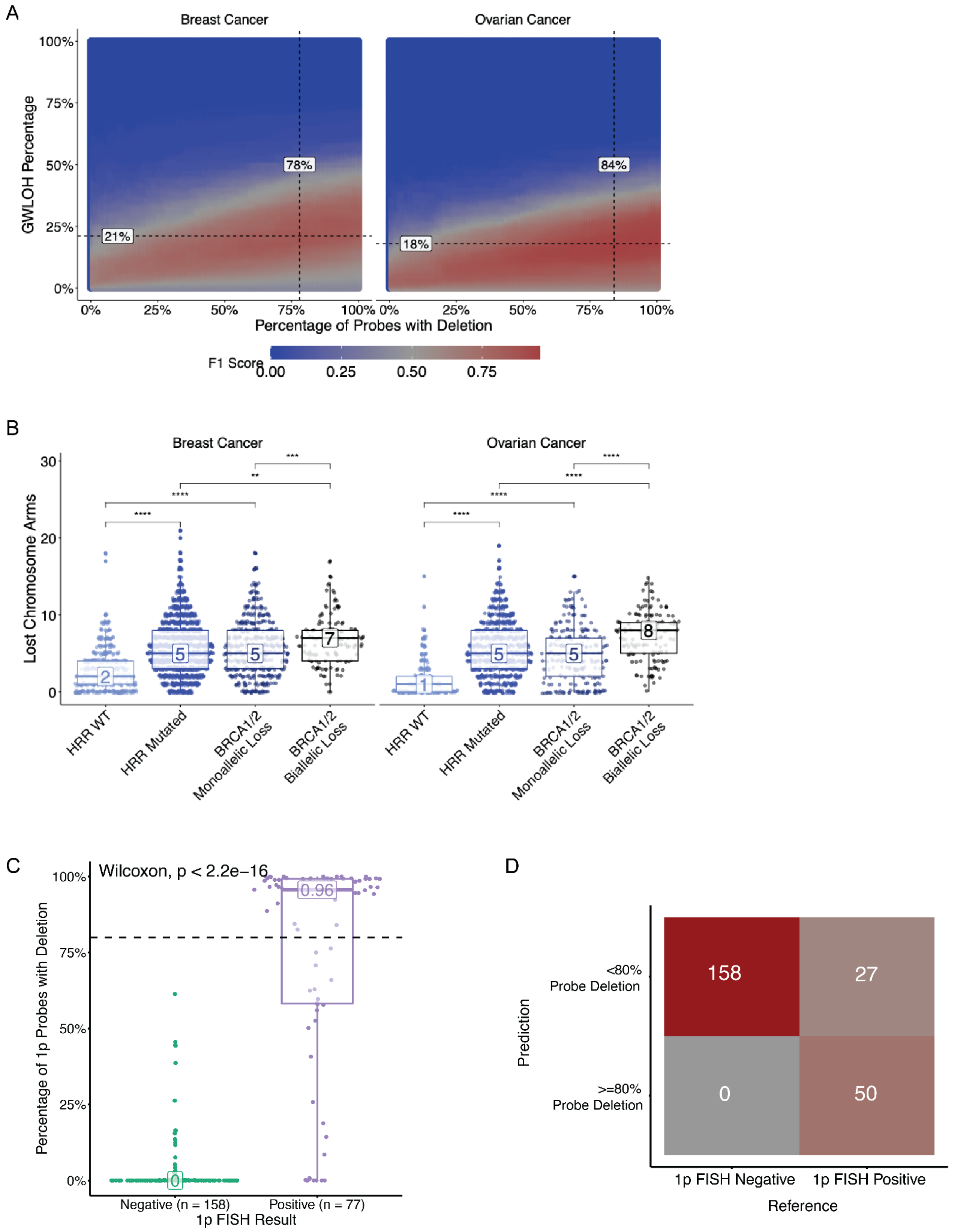


**Supplemental Figure 2. HRD-DNA captures chromosome arm losses.** (A) Relationship between percentage of probes with deletion (x-axis), gwLOH threshold for HRD+ samples, and resulting F1-score (color). Dashed lines represent the maximum F1-score for each cancer cohort in the training samples. (B) For both breast and ovarian cancer, samples with HRR mutations or *BRCA1/2* monoallelic loss have more lost chromosome arms than HRR-WT samples (Wilcoxon test, (** p-value < 0.01, *** p-value < 0.001, **** p-value < 0.0001). Statistical differences between HRR-WT and BRCA biallelic loss were not shown, but all were significant (p-value < 0.0001 for Wilcoxon test). (C) Glioma samples with external FISH results were used to calculate the accuracy of the selected probe loss threshold. Samples with a positive FISH result have a higher percentage of 1p probes with deletion (Wilcoxon test). (D) Confusion matrix for gliomas samples with external 1p FISH testing compared to predicted 1p loss based on an 80% probe loss threshold, indicating 89% concordance.

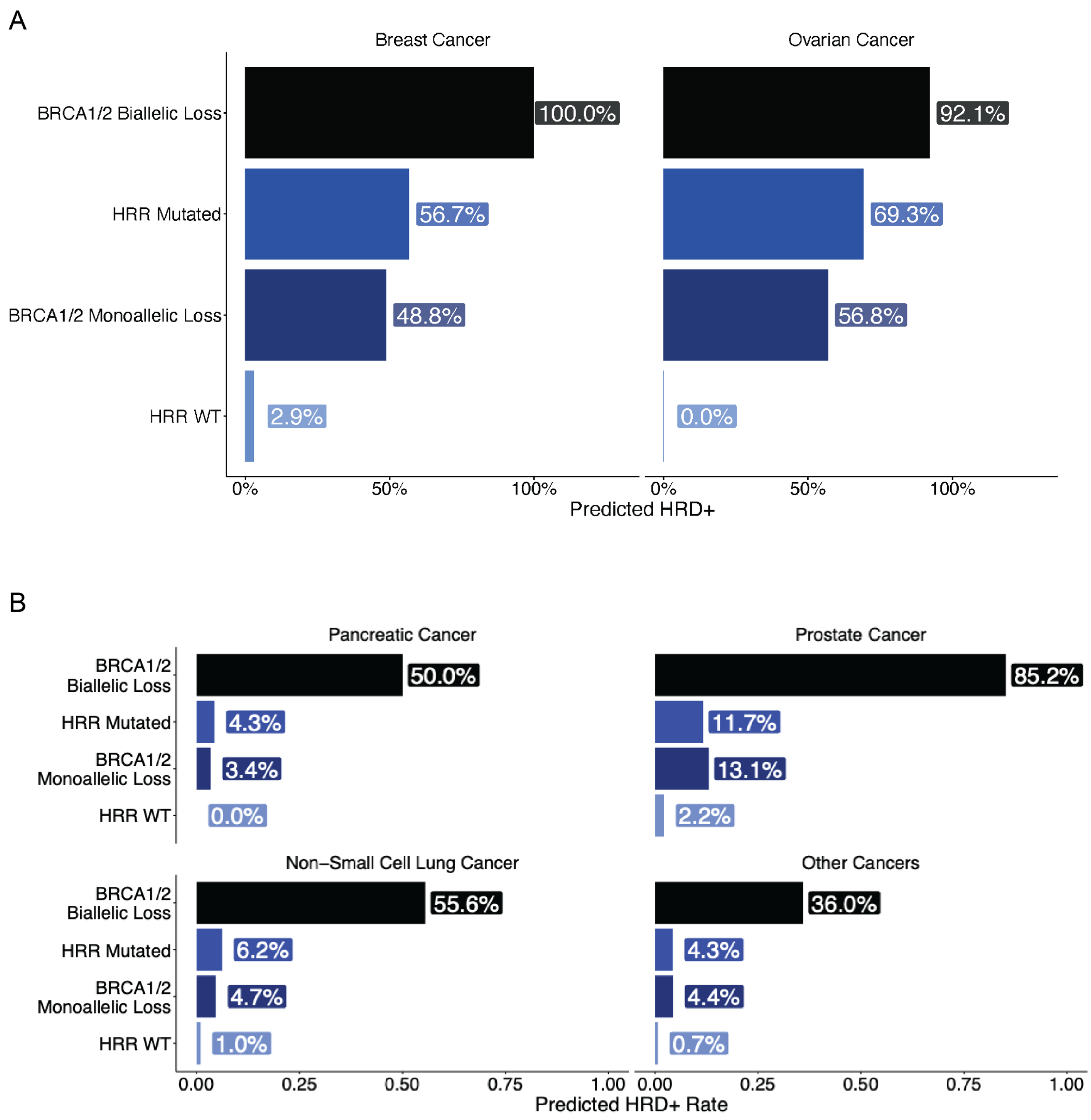


**Supplemental Figure 3. Rates of HRD+ predictions differ by HRR genotype.** The rate of HRD+ predictions was compared within the evaluation cohort and stratified by mutation status for the HRD-DNA (A) and HRD-RNA (B) models. Both models are enriched for HRD+ predictions in the HRR Mutated and *BRCA1/2* monoallelic loss sample sets when compared to the HRR-WT sample set (Fisher's exact test, p-value < 0.0001).

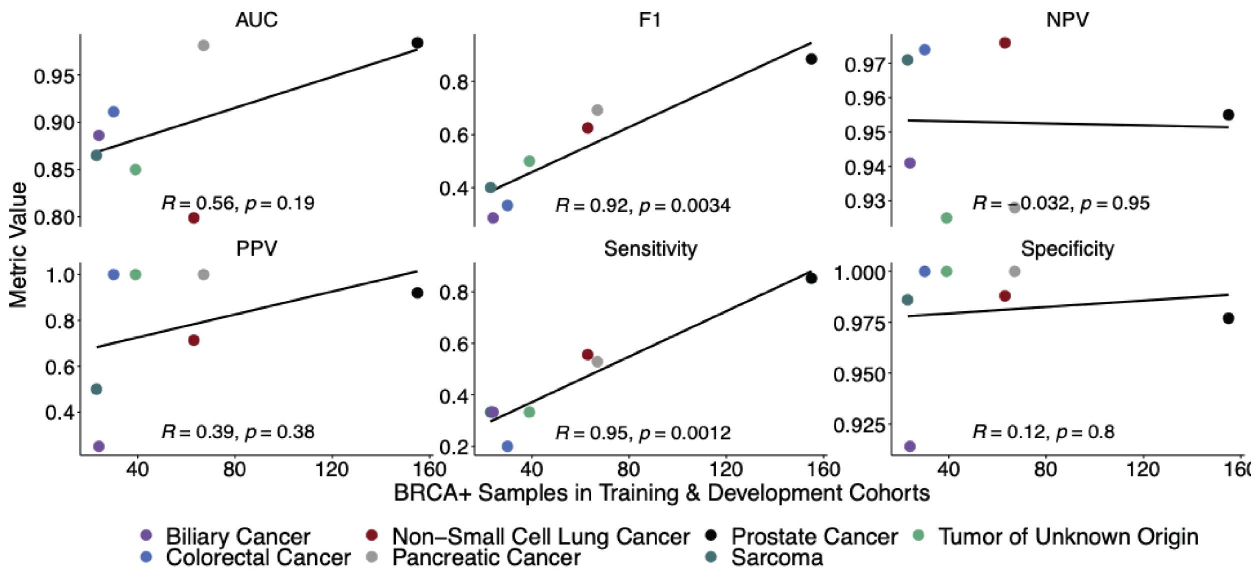


**Supplemental Figure 4. Correlation between BRCA+ samples in training and performance metrics on a cancer cohort-level.** The number of samples with *BRCA1/2*-biallelic loss in the training and development sets is positively correlated with the F1 score and sensitivity of the model on the evaluation cohort, suggesting a biological and/or modeling constraint in cancer types with a low prevalence of BRCA deficiency.

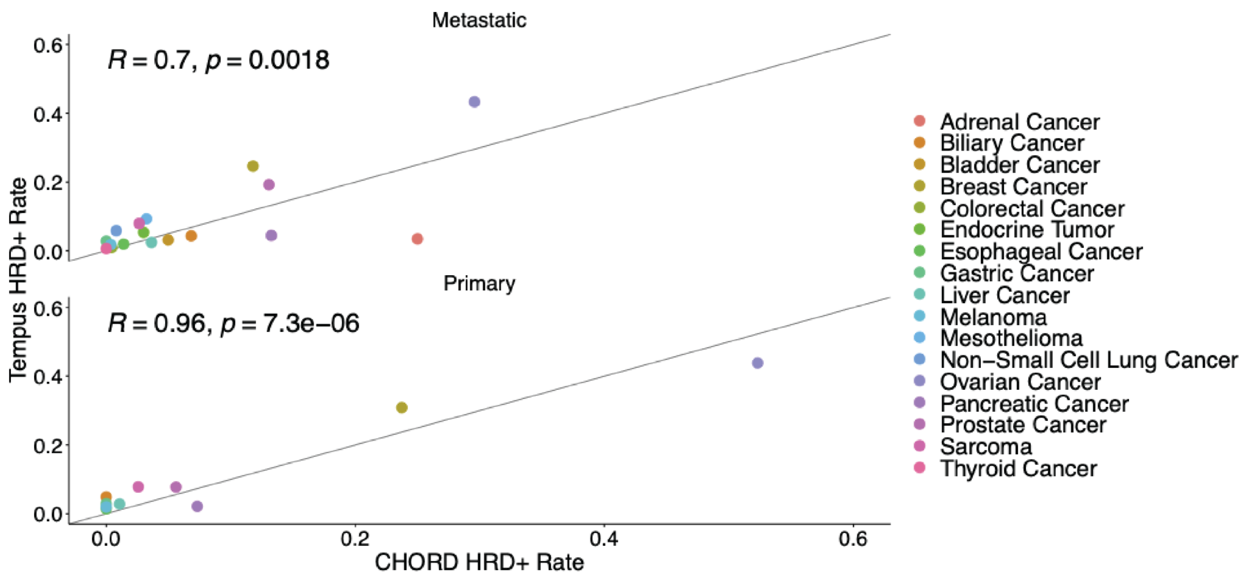


**Supplemental Figure 5. Cancer types HRD+ prevalence stratified by primary and metastatic in CHORD vs Tempus.** Predicted HRD+ rates within cancer types are highly correlated between CHORD and Tempus, with primary cancers showing a stronger correlation than metastatic cancers. The solid line represents equal predicted frequencies of HRD between CHORD and Tempus.

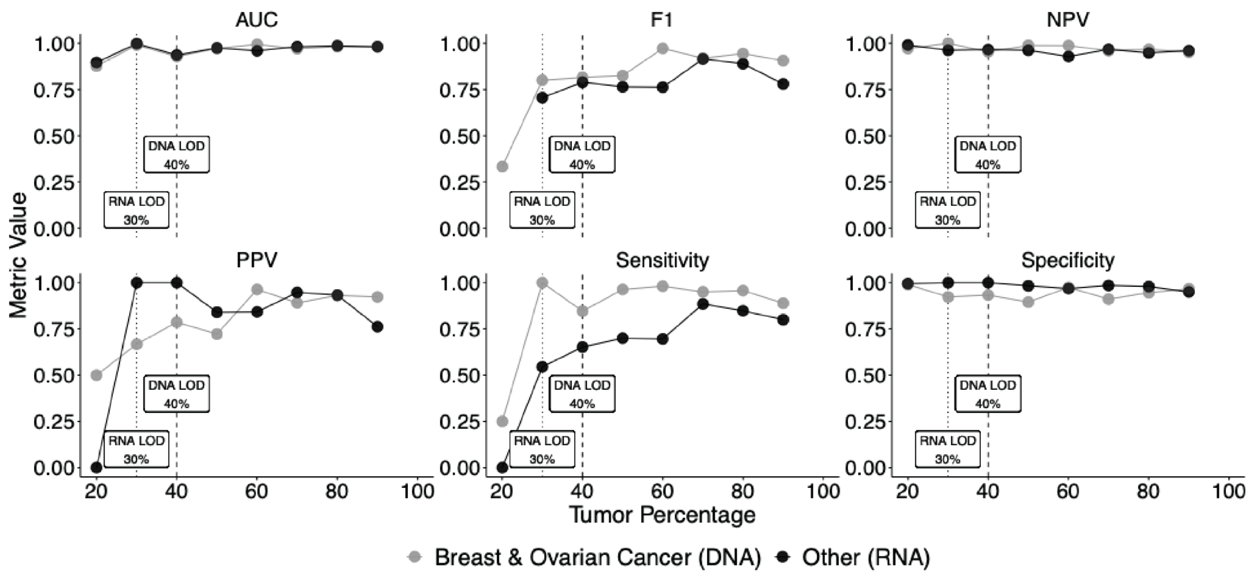


**Supplemental Figure 6. Performance metrics across different tumor percentages to determine the limit of detection of HRD-DNA and HRD-RNA.** The performance of HRD-DNA and HRD-RNA on the evaluation cohort was stratified by pathologist-estimated tumor percentage. The limit of detection (LOD) was determined by the minimum tumor purity that predicts BRCA biallelic samples from HRR-WT samples with 70% PPV, resulting in an HRD-DNA LOD of 40% and an HRD-RNA LOD of 30%.

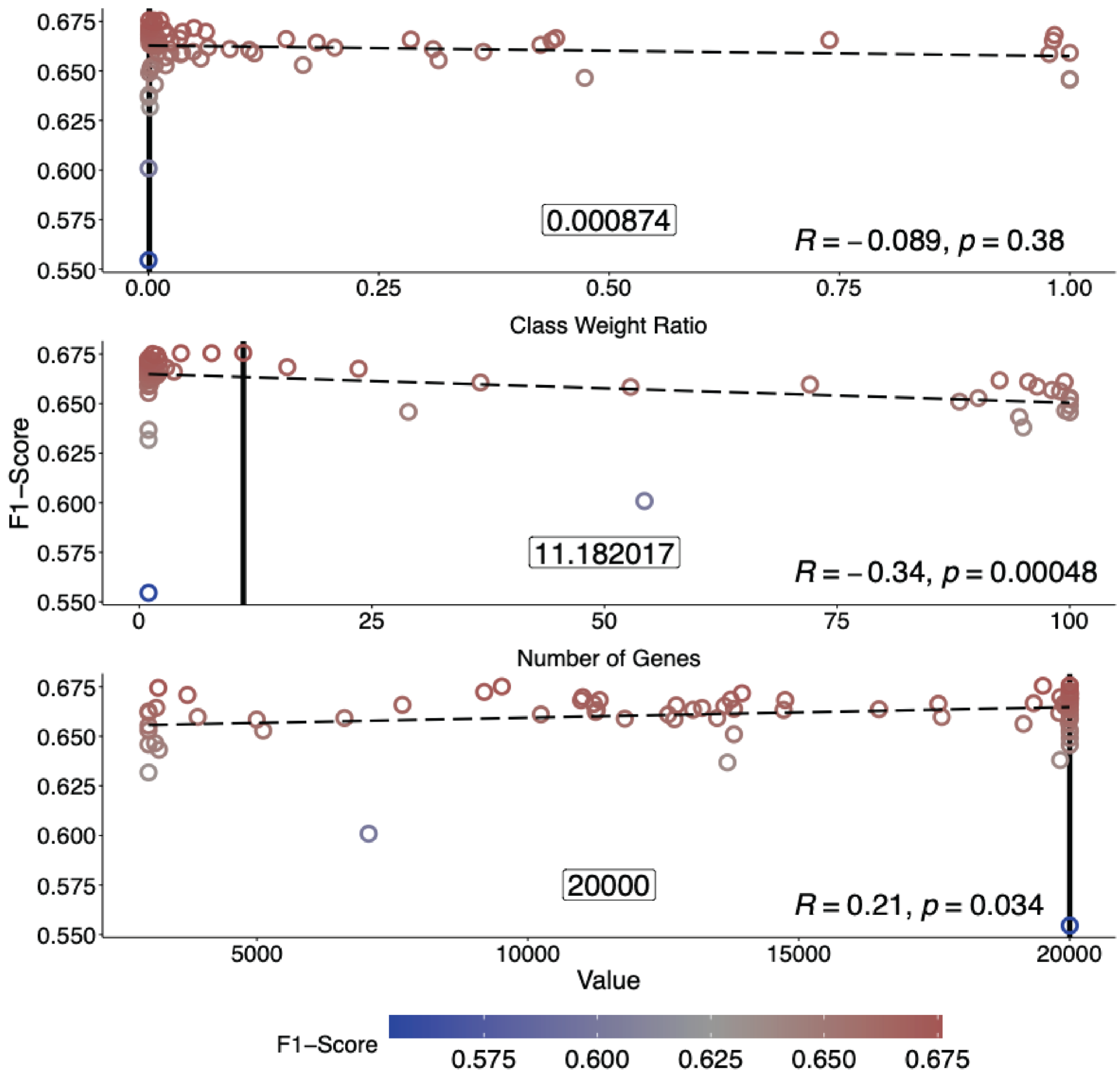


**Supplemental Figure 7. Parameter tuning for the HRD-RNA model.** Three hyperparameters were tuned with Bayesian optimization to achieve optimal performance of the HRD-RNA model, where optimal performance was defined as the maximum F1-score on the development samples (Methods). Optimization identified a significant correlation between both the class weight ratio and performance and the number of genes and performance.

| Biopsy Location | Positives | Sensitivity | Specificity | PPV | NPV | F1 | AUC |
|---|---|---|---|---|---|---|---|
| Breast & Ovarian Cancer | | | | | | | |
| Metastatic | 36 | 0.972 | 0.944 | 0.972 | 0.944 | 0.972 | 0.997 |
| Primary | 19 | 0.947 | 1.000 | 1.000 | 0.917 | 0.973 | 0.986 |
| Indeterminate - Missing Data | 8 | 0.875 | 1.000 | 1.000 | 0.917 | 0.933 | 0.966 |
| Indeterminate - Lymph Involvement | 4 | 1.000 | 1.000 | 1.000 | 1.000 | 1.000 | 1.000 |
| Other Cancers | | | | | | | |
| Metastatic | 29 | 0.448 | 0.994 | 0.867 | 0.953 | 0.591 | 0.953 |
| Primary | 29 | 0.690 | 0.990 | 0.800 | 0.982 | 0.741 | 0.956 |
| Indeterminate - Missing Data | 9 | 0.667 | 0.985 | 0.667 | 0.985 | 0.667 | 0.874 |
| Indeterminate - Lymph Involvement | 11 | 0.636 | 1.000 | 1.000 | 0.960 | 0.778 | 0.940 |

**Supplemental Table 1. Performance of HRD-DNA and HRD-RNA by biopsy tissue site.** HRD-DNA (breast & ovarian cancer) has robust performance across biopsy settings, demonstrating stable sensitivity and specificity across metastatic, regional lymph node, and primary biopsies. HRD-RNA (other cancers) has high specificity across all biopsy settings. Differences in sensitivity and F1 score may be influenced by the representation number of each cancer cohort in the respective biopsy site.
